# Cytokine profile of saliva in adults with focal enamel demineralization after orthodontic treatment

**DOI:** 10.1101/2025.11.19.25340370

**Authors:** A.A. Bayakhmetova, A.O. Seydekhanova, A.N. Primbayeva, A.M. Duisenbayev

## Abstract

**Background and aim:** the saliva content and correlation with the intensity of caries of proinflammatory cytokines IL-1ß, IL-6, IL-8 and anti-inflammatory cytokines IL-4 and IL-10 were studied in patients with FED after long-term orthodontic treatment.

**Materials and methods:** 26 patients aged 18 to 30 years were divided into three groups: a group of patients without caries, a group with compensated caries with a caries intensity index (DMF decayed, missing, filled) value of less than 5.0 and a group with decompensated caries with a caries intensity index value of more than 5.0.

**Results:** based on the results obtained, it was found that patients with FED have violations both in the system of nonspecific and specific local immunity, which is more pronounced in patients with decompensated caries. The essential role of the proinflammatory cytokine IL-6 was noted, the content of which in saliva significantly prevails in patients with decompensated caries.

**Conclusion:** the study of the correlation relationships of the studied cytokines allowed us to determine the significant role of IL-6 and IL-4 cytokines and their ratios in the pathogenesis of FED.

## Introduction

Focal enamel demineralization (FED) is one of the frequent complications of long-term non-removable orthodontic treatment using braces. The development of FED is associated with an increase in microbial colonization and the failure of colonization resistance of the oral cavity, an irrational diet, a low level of oral hygiene, errors in orthodontic treatment and its duration ^1,2,3^. With FED, not only quantitative, but also qualitative changes in the oral microbiome occur with the prevalence of Str. mutans, Lactobacillus spp. and potentially pathogenic gram-negative bacteria. In the initiation of the FED, the main role of Str. mutans is emphasized ^3,4,5^.

Long-term non-removable orthodontic treatment is an extreme factor that violates the homeostasis of the oral cavity and requires adequate and effective participation of the immune system to normalize the microbiome of the oral cavity. Dysbiosis and inferiority of protective factors of saliva determine the susceptibility of enamel to pathogenic factors. Saliva is considered as a natural primary protective system for the surfaces of teeth, the study of some parameters of which reflects the relationship with caries ^6,7,8,9,10,11,12^. Saliva plays an important role in maintaining balance and symbiotic relationships between the body and the oral microbiota. The presence of many factors of nonspecific and specific immunity in saliva requires adequate regulation of their interaction. To a large extent, this is determined by regulators, or mediators of immune reactions, which include cytokines. Cytokines function in a complex interconnected cytokine network in which the secretion of one cytokine leads to the appearance and activation of others. These properties of cytokines provide extensive functionality with participation not only in the regulation of systemic immune reactions, but also in the implementation of local immunity factors ^13,14^.

Cytokines are synthesized by lymphocytes, monocytes, macrophages, granulocytes and other cells in response to microbial invasion, antigenic irritation or tissue damage. Characteristic are the extreme promptness of synthesis, high activity in negligible amounts, polyfunctionality or pleiotropy. These properties of cytokines provide high efficiency and reliability of their biological action ^14,15,16^. There are pro- and anti-inflammatory cytokines. Pro-inflammatory cytokines are represented by interleukins 1,2,6,8, TNF-α, interferon-γ, and interleukins 4,10, TGF-β are classified as anti-inflammatory cytokines. Cytokines play a decisive role in the nature of the emerging adaptive immune response ^17,18^. Local protective immune reactions in the oral cavity, such as phagocyte chemotaxis, phagocytosis and antibody formation, the formation of cellular or humoral immunity are regulated by cytokines.

Immunological examination of patients aged 18 to 45 years with caries revealed a significant increase in the content of proinflammatory cytokine IL-1ß in saliva, while the concentration of secretory immunoglobulin A (sIgA) and IFN-γ decreased. The authors named these changes as predisposing factors to the development of caries ^19^. However, studies devoted to the study of the cytokine profile of saliva of adult patients with caries are rare. Basically, the literature data on the cytokine profile of saliva relate to caries of childhood and adolescence and show a statistically significant increase in the level of proinflammatory cytokines IL-6, IL-8 and TNF-α in comparison with similar indicators in healthy people ^20,21,22^.

The role of proinflammatory cytokines in the pathogenesis of demineralization of hard tooth tissues is not sufficiently clear, however, it may reflect the nature of the emerging local adaptive immune response and the level of local immunity in the oral cavity.

The aim of our study was to study the cytokine profile of mixed unstimulated saliva in adult patients with FED after long-term orthodontic treatment.

## Material and methods

26 people (5 men and 21 women) aged from 18 to 20 years (10 people) and 20-29 years (16 people) who received long-term fixed orthodontic treatment were examined. Informed consent for examination and treatment was received from all patients. The duration of non-removable orthodontic treatment in the form of a bracket system in all cases of observation was more than one year. The criteria for selecting patients were the absence of somatic pathology, age from 18 to 30 years, long-term fixed orthodontic treatment for at least 1 year. After the removal of the bracket system, FED was diagnosed in 24 patients. Two observation groups were formed depending on the known indicator of the intensity of caries of the DMF. The groups with FED were represented by 10 patients with a DMF of no more than 5.0, and 14 people with a DMF of more than 5.0. The comparison group consisted of 10 practically healthy people under the age of 30 and with no caries. In all examined patients on an empty stomach, mixed unstimulated saliva was collected by spitting into a sterile tube for 10 minutes in the morning from 9 to 11 o’clock. Saliva was centrifuged at 10000 r.p.m, the prepared samples were stored at a temperature of – 20□. To determine the level of cytokines in saliva samples, a “sandwich” version of solid-phase enzyme immunoassay was used using mono- and polyclonal antibodies to cytokines IL-1ß, IL-4, IL-6, IL-8 and IL-10 using a set of reagents and instructions from Vector Best (Russia). When choosing cytokines for the study, we were guided by the literature data. Sandwich-ELISA, consists in the following: one type of monoclonal antibodies to a certain cytokine is immobilized on the inner surface of the cells of the tablets for research. The test material and the corresponding standards and controls are introduced into the wells of the tablet. After incubation and washing, the second monoclonal antibodies to another epitope of this cytokine, conjugated with an indicator enzyme (horseradish peroxidase), are introduced into the wells. After incubation and washing, a substrate is introduced into the cells-hydrogen peroxide with chromogen. During the enzymatic reaction, the intensity of the color of the wells changes, which is measured on an automatic photometer for tablets.

Statistical analysis of the obtained digital material was carried out in the SPSS Statistics version 22 program using descriptive statistics, the Mann-Whitney criterion for comparing the results obtained in groups and correlation analysis with the determination of Spearman’s rank correlation coefficient.

## RESULTS

When studying the concentration in the mixed unstimulated saliva of patients with FED with compensated caries, there was a significant decrease in the content of pro-inflammatory cytokines IL-6 and IL-8 (P<0.002, P<0.05) and anti-inflammatory cytokine IL-4 (P<0.002) in saliva. The index of IL-6 content in accordance with Spearman’s rank correlation coefficient had a significant direct correlation with the concentration of the proinflammatory cytokine IL-8 in saliva (r=0.64, P<0.05). A direct correlation was found between the indicators of the content of IL-8 and IL-10 in saliva (r=0.69, P <0.05).

With decompensated caries, patients with FED retained a decrease in the concentration of IL-8 in saliva (P<0.05), and the content of IL-6 and IL-4 increased and was significantly higher than with compensated caries (P<0.001, P<0.02). Between the indicators of the content of IL-6 and IL-4 in saliva a direct correlation was revealed (r=0.61, P <0.05).

To clarify the nature of the relationship between pro- and anti-inflammatory cytokines, we conducted a study of the ratio coefficients between pro-inflammatory cytokines I□-1β, I□-6, I□-8 and anti-inflammatory cytokines I□-4 and I□-10 in the examined groups. The nature of these ratios allows us to judge the relative deficiency or hyperproduction of a particular cytokine. The results obtained are shown in table 2 and in diagram 2.

**Table 1.**
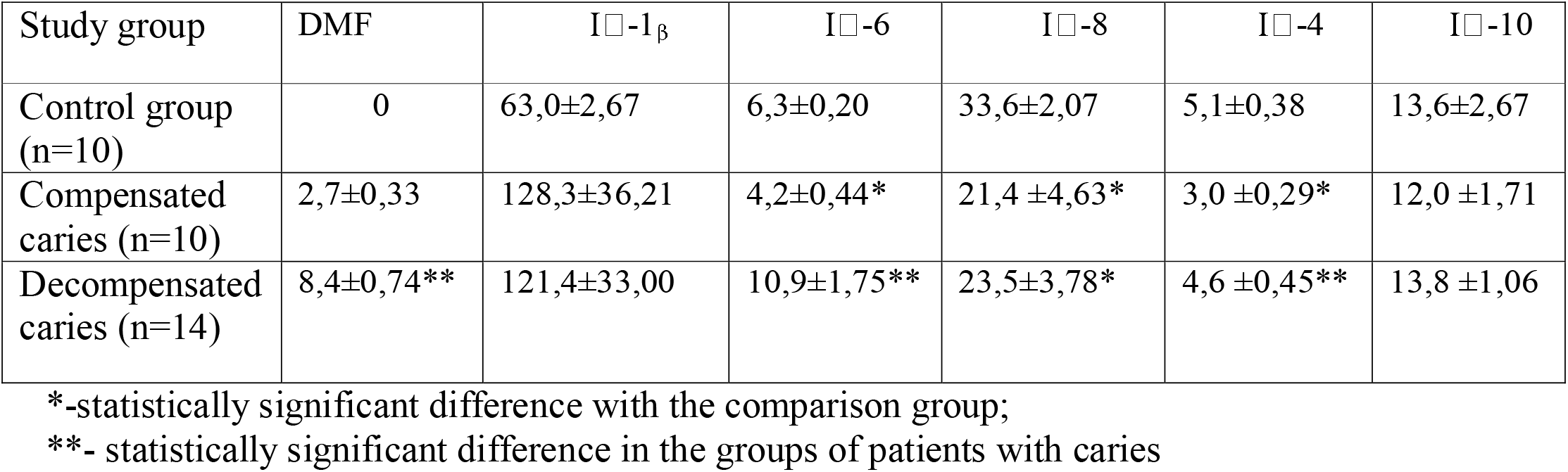
Cytokine profile of mixed unstimulated saliva in patients with FED with different caries intensity (pg/ml).

**Table 2.** The ratio of pro-inflammatory and anti-inflammatory cytokines in patients with FED with different caries intensity.

| Study group | DMF | I□-1<sub>β</sub>/ I□-4 | I□-1<sub>β</sub>/ I□-1 | I□-6/ I□-4 | I□-6/ I□-10 | I□-8/ I□-4 | I□-8/ I□-10 |
| --- | --- | --- | --- | --- | --- | --- | --- |
| Control group (n=10) | 0 | 13,4±1,84 | 7,8±2,19 | 1,3±0,16 | 0,9±0,27 | 7,1±0,99 | 4,3±1,31 |
| Compensated caries (n=10) | 2,7±0,33 | 47,5±12,79 | 11,1±3,70 | 1,4±0,20 | 0,4±0,05 | 8,2±2,18 | 1,8±0,33* |
| Decompensated caries (n=14) | 8,4±0,74** | 34,3±11,07 | 13,0±6,44 | 2,4±0,28*,** | 0,9±0,13** | 6,0±1,29 | 1,9±0,29 |

\*-statistically significant difference with the comparison group;
\*\* - statistically significant difference in the groups of patients with caries

**Diagram 1.**
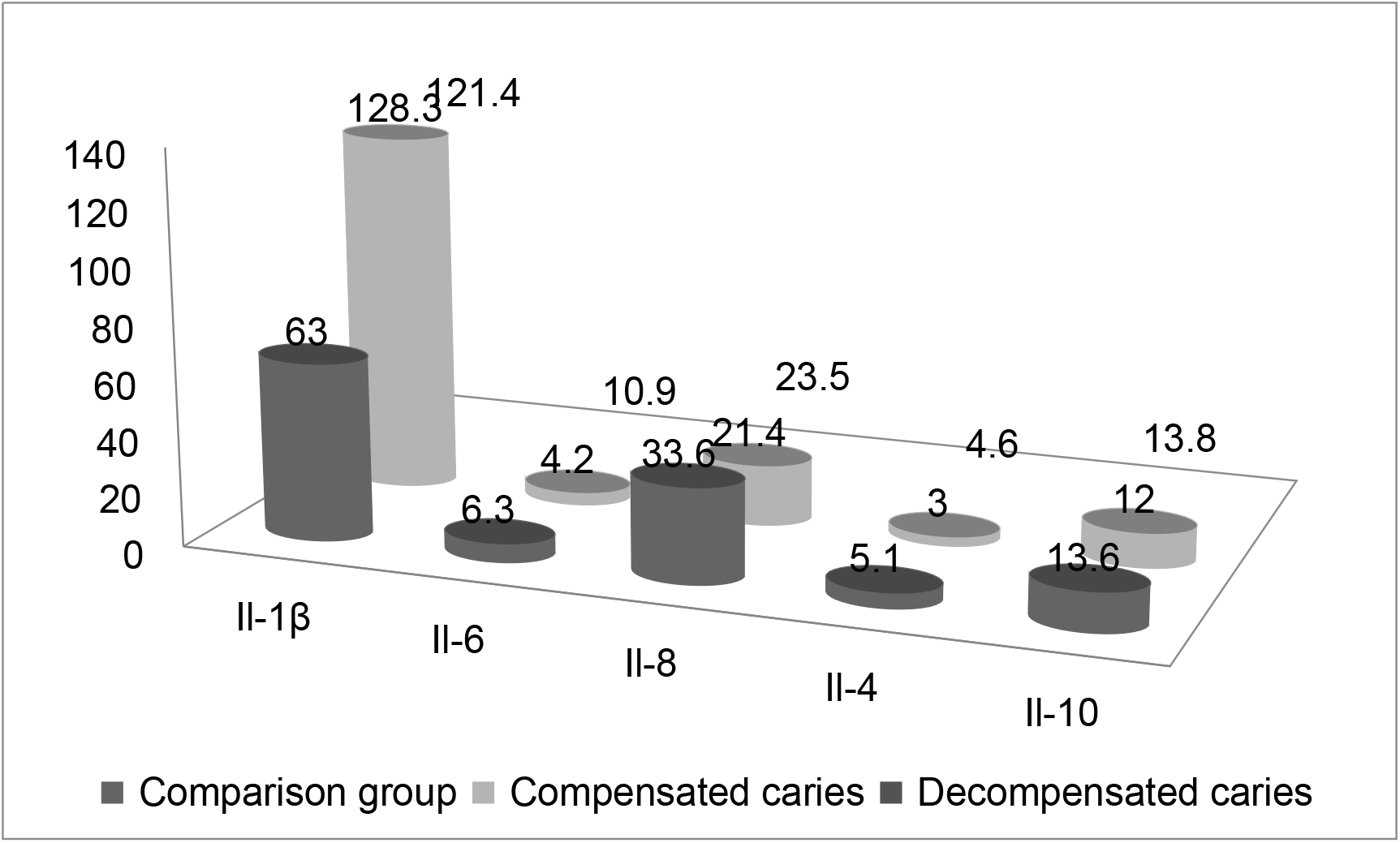
Cytokine profile of mixed unstimulated saliva in patients with FED with different caries intensity.

**Diagram 2.**
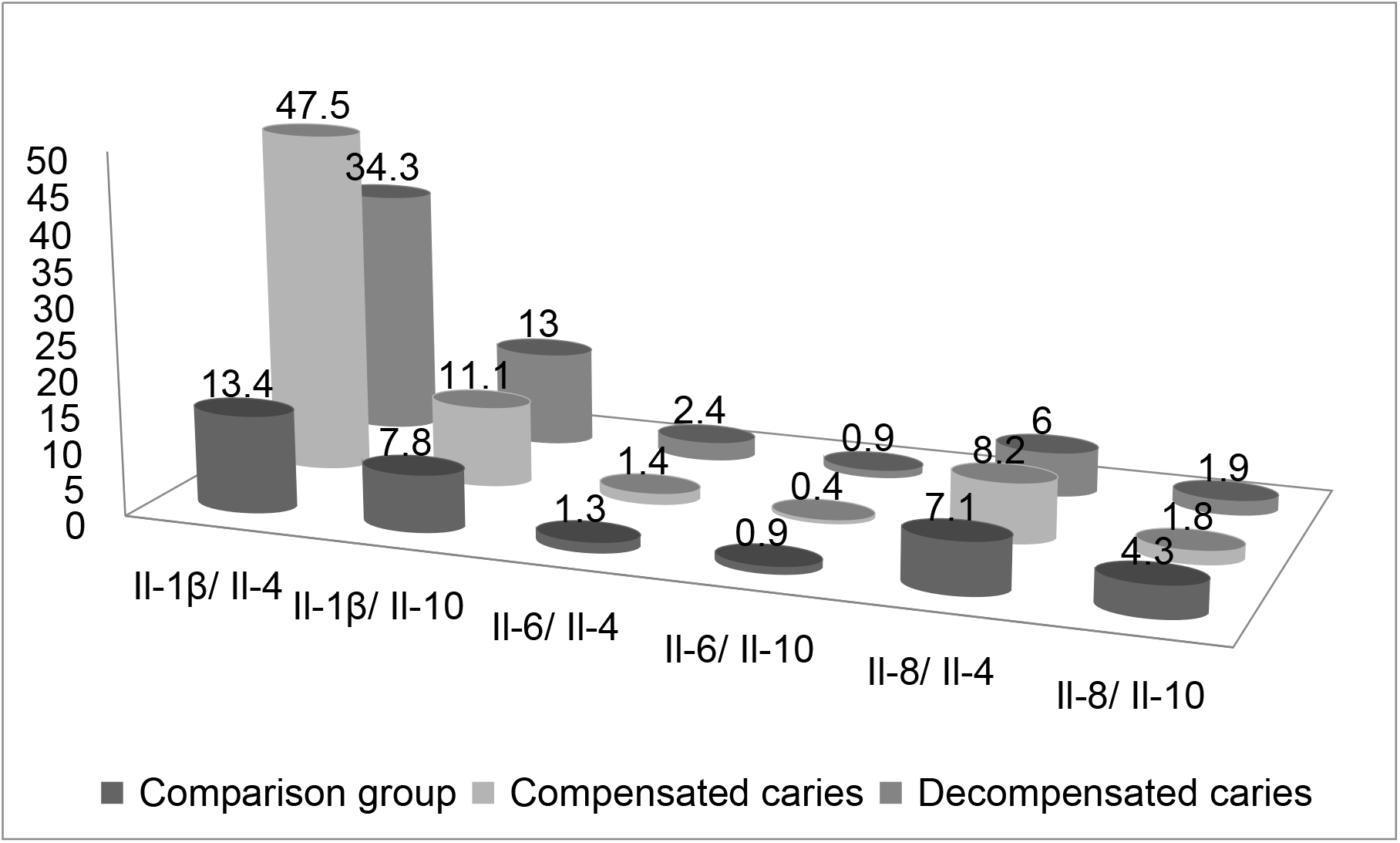
Ratios of proinflammatory and anti-inflammatory cytokines in patients with FED with different caries intensity.

As follows from the results shown in Table 2, the ratio I□-1β/I□-4 reflected an increase in the content of proinflammatory cytokine I□-1β in saliva, which was expressed in an unreliable increase in this ratio with compensated and decompensated caries. The ratios I□-1β/ I□-10 in the examined groups also did not show a significant difference from similar indicators of the comparison group.

The changes in the ratio I□-6/I□-4 were different. In the group with decompensated caries, its value was significantly higher not only than in the comparison group (P<0.005), but also in patients with compensated caries (P<0.03). With compensated caries, when compared with the group with decompensated caries, a significant decrease in the average value of the ratio I□-6/ I□-10 (P<0.05) and a decrease in the ratio I□-8/ I□-10 when compared with the comparison group (P<0.05) was found.

## DISCUSSION

The primary etiological factors in caries are microbial biofilm, a highly carbohydrate diet of soft and sticky consistency, as well as a low level of oral hygiene. Defects of local immunity play a significant role in the pathogenesis of caries ^6,7^. The local immunity of the oral cavity is represented by a variety of nonspecific and specific protective factors of saliva, providing effective protection against cariesogenic factors of microbial biofilm ^13^.

Some pro-inflammatory and anti-inflammatory cytokines (IL-6, IL-8, TNF-α, IL-1ß, IL-4, IL-10) are associated with the pathogenesis of caries ^13^. The anti-inflammatory cytokine IL-4 is synthesized by mast cells, T-helper cells of the second type, eosinophils and basophils. IL-4 stimulates the formation of reparative macrophages of the second type, promotes the differentiation of naive T-lymphocytes into T-helper cells of the second type and their transformation into plasma cells, which leads to a humoral pathway of immunity with the synthesis of specific antibodies. The anti-inflammatory cytokine IL-10 is produced by macrophages and regulatory T cells, inhibits the functions of monocytes/macrophages, the secretion of pro-inflammatory cytokines IL-1,6,8,12, TNF-α, IFN-γ by various cells. The lack of anti-inflammatory cytokine IL-10 indicates a lack of nonspecific immunity, pro-inflammatory cytokine IL-8 indicates inhibition of phagocytic activity with impaired chemokinesis. However, an adequate assessment of the cytokine profile can be carried out with mandatory consideration of the ratios of pro- and anti-inflammatory cytokines in a complex interconnected cytokine network, in which the secretion of one cytokine leads to the appearance and activation of others ^23^. Cytokine IL-6 is a pro- and anti-inflammatory cytokine that is synthesized by macrophages, monocytes, fibroblasts and endothelial cells. It can inhibit the synthesis of activated macrophages and T-lymphocytes of the proinflammatory cytokine TNF-α and IL-1ß. A correlation was established between the increased level of IL-6 in saliva and the intensity of caries. IL-1ß is a multifunctional cytokine with a wide spectrum of action, synthesized by macrophages and monocytes, plays a key role in the development and regulation of nonspecific protection and specific immunity, one of the first to be included in the body’s protective response under the action of pathogenic factors.

Our results show that patients with FED with compensated caries after prolonged non-removable orthodontic treatment are likely to have violations of the formation of adaptive local immunity of the humoral type due to a significant decrease in the content of Il-4 in saliva. A decrease in the concentration in the saliva of patients with FED II-8 leads to an imbalance of non-specific immunity, insufficient phagocytic activity of phagocytic cells. With decompensated caries, patients with FED have a significant voltage associated with an increase in IL-6 synthesis, which is manifested by a significant increase in the digital values of the ratios of IL-6/ and IL-4, and IL-6/ and IL-10.

## CONCLUSION

The ratio of pro-inflammatory and anti-inflammatory cytokines is a crucial moment in the outcome of inflammation of the soft tissues of the oral cavity, but the participation of cytokines in the pathogenesis of demineralization of hard tooth tissues is not adequately studied. However, it is obvious that in the pathogenesis of an infectious disease, which is caries, the role of local oral immunity and cytokines as immune regulators cannot be overestimated. Thus, the results of studying the indicators of local immunity, namely the cytokine profile of mixed unstimulated saliva of patients with FED, allowed us to come to the following conclusions.

In patients with FED, there is a tension of local immunity, which is manifested by an imbalance of the cytokine profile of mixed unstimulated saliva with a predominance of the content of proinflammatory cytokines IL-1β, IL-6, with a decrease in the concentration of proinflammatory cytokine IL-8 and anti-inflammatory cytokine IL-4.

With decompensated caries in patients with FED, the content of pro-inflammatory cytokine IL-6 and anti-inflammatory cytokine IL-4, the ratio of IL-6/IL-4 were significantly higher than similar indicators in patients with compensated caries.

The obtained results suggest the key role of the pro-inflammatory cytokine IL-6 and the failure of the anti-inflammatory cytokines IL-4 and IL-10 in the formation of a local immune response in decompensated caries.

The study of correlational relationships of the studied cytokines allowed us to determine the significant role of IL-6 and IL-4 cytokines in the pathogenesis of FED.

Our results and conclusions are based on a small number of observations and require further research.

## Data Availability

All data produced in the present study are available upon reasonable request to the authors
All data produced in the present work are contained in the manuscript

